# ConceptWAS: a high-throughput method for early identification of COVID-19 presenting symptoms

**DOI:** 10.1101/2020.11.06.20227165

**Authors:** Juan Zhao, Monika E Grabowska, Vern Eric Kerchberger, Joshua C. Smith, H. Nur Eken, QiPing Feng, Josh F. Peterson, S. Trent Rosenbloom, Kevin B. Johnson, Wei-Qi Wei

## Abstract

**Objective:** Identifying symptoms highly specific to COVID-19 would improve the clinical and public health response to infectious outbreaks. Here, we describe a high-throughput approach – Concept-Wide Association Study (ConceptWAS) that systematically scans a disease’s clinical manifestations from clinical notes. We used this method to identify symptoms specific to COVID-19 early in the course of the pandemic.

**Methods:** Using the Vanderbilt University Medical Center (VUMC) EHR, we parsed clinical notes through a natural language processing pipeline to extract clinical concepts. We examined the difference in concepts derived from the notes of COVID-19-positive and COVID-19-negative patients on the PCR testing date. We performed ConceptWAS using the cumulative data every two weeks for early identifying specific COVID-19 symptoms.

**Results:** We processed 87,753 notes 19,692 patients (1,483 COVID-19-positive) subjected to COVID-19 PCR testing between March 8, 2020, and May 27, 2020. We found 68 clinical concepts significantly associated with COVID-19. We identified symptoms associated with increasing risk of COVID-19, including “absent sense of smell” (odds ratio [OR] = 4.97, 95% confidence interval [CI] = 3.21–7.50), “fever” (OR = 1.43, 95% CI = 1.28–1.59), “with cough fever” (OR = 2.29, 95% CI = 1.75–2.96), and “ageusia” (OR = 5.18, 95% CI = 3.02–8.58). Using ConceptWAS, we were able to detect loss sense of smell or taste three weeks prior to their inclusion as symptoms of the disease by the Centers for Disease Control and Prevention (CDC).

**Conclusion:** ConceptWAS is a high-throughput approach for exploring specific symptoms of a disease like COVID-19, with a promise for enabling EHR-powered early disease manifestations identification.

## Introduction

As of October 14, 2020, over 7.7 million people in the United States (U.S.) and 37 million worldwide were infected with coronavirus SARS-CoV-2, the agent responsible for COVID-19 [1]. The virus’s high transmissibility, lack of native immunity, and the dearth of effective treatments make managing COVID-19 uniquely challenging. Hence, early recognition of specific COVID-19 symptoms plays an essential role in the clinical and public health response, enabling rapid symptom screening, diagnostic testing, and contact tracing.

Early in the pandemic, physicians observed fever, cough, and shortness of breath as presenting symptoms of COVID-19; however, these symptoms are common to many viral and bacterial illnesses [2]. Subsequently, as new symptoms were reported, health departments and ministries updated the list of COVID-19 symptoms; for example, the U.S. CDC and the Department of Health and Social Care in the UK added loss of smell or taste, a highly indicative symptom [3] to the list in late April and mid-May, respectively [4,5]. Therefore, methods of identifying the specific COVID-19 symptoms early during a pandemic are required, which is crucial to inform the public on when to present for testing, and potentially reduce the ultimate size of the outbreak, lowering overall morbidity and mortality.

Recent efforts to track COVID-19 symptoms have used methods such as scanning publications or twitter [6,7], deploying questionnaires [8], or releasing apps to self-report symptoms [9]. However, results from publications and questionnaires may be delayed; data from social media or self-reported apps do not always include proper controls and lack physiological assessments to determine the COVID-19 status. Electronic Health Records (EHR) data has also been used to characterize COVID-19, due to the availability of routinely collected medical data. However, existing studies were limited by structured data (e.g., coded diagnoses, procedures, or lab tests) [10,11] and lacked a portable and high-throughput approach [12].

Here, we present a high-throughput approach (ConceptWAS) for early identification of clinical manifestations of COVID-19 using natural language processing (NLP) on EHR clinical notes. By examining the EHR-derived concepts related to patients’ signs or symptoms, ConceptWAS assesses whether any of the concepts are associated with the presence or absence of a disease (e.g., COVID-19). Using this approach, we identified the symptoms that were specific to COVID-19. In particular, we performed ConceptWAS using the cumulative data every two weeks to demonstrate the timeline of emerging symptoms. We also conducted a chart review to validate the significant associations.

## Methods

### Study setting

The study was performed at Vanderbilt University Medical Center (VUMC), one of the largest primary care and referral health systems serving over one million patients annually from middle Tennessee and the Southeast United States. We used data from patients represented in the VUMC EHR aged ≥18 years. The study was approved by the VUMC Institutional Review Board (IRB #200512).

### Cohort definition

We firstly identified patients who received at least one SARS-CoV-2 polymerase chain reaction (PCR) test between March 8 (when the first COVID-19 emerged in VUMC) and May 27, 2020 (Figure A.1). The COVID-19 status was determined using the PCR test result. The case group (COVID-19-positive) was defined as patients who had >=1 PCR positive result, and the control group (COVID-19-negative) consisted of patients with only negative PCR tests. We excluded patients who had no clinical notes on the day the PCR test was ordered.

### Data collection

We extracted clinical notes from 24 hours prior to PCR testing date (day_0_) for COVID-19-positive and negative patients (>86% of patients had at least one note within the time window, see Figure B.1). If a patient first tested negative and then subsequently tested positive or if a patient were tested positive more than once, we used the date of the first positive PCR test as day_0_. We also segmented the study period into a two-week interval and used the cumulative data every two weeks to perform a temporal analysis.

### Concept extraction

We used KnowledgeMap Concept Indexer (KMCI [13]) to preprocess the clinical notes and extract concepts (Figure C.1). KMCI is a local NLP pipeline developed at VUMC for medical notes processing and entity recognition, which has been used for several clinical and genomic studies [13–15]. The preprocessing includes sentence boundary detection, tokenization, part-of-speech tagging, section header identification. The concepts were represented as Unified Medical Language System concept unique identifiers (UMLS CUIs). Since we focused on capturing clinical manifestations of COVID-19, we restricted the concepts to SNOMED Clinical Terms and specific semantic types, e.g., finding, sign or symptom, disease or syndrome, individual behaviors, or mental process (see full list in Table C.1).

### Assertion and negation detection

A main challenge of clinical NLP is to accurately detect the clinical entities’ assertion modifier such as negated, uncertain, and hypothetical (e.g. describe a future hypothetical or instruction for patients). We took the following steps to post-process the KMCI output to remove CUIs that were uncertain, and hypothetical. We first excluded any concepts that arose from family history sections. Next, we removed any sentences with future sense or subjunctive mood *(e*.*g. “should”, “could”*, or *“if”*) that describe a hypothetical or instruction for patients. We excluded inquiry sentences that served as the template questions without a simple confirmed answer (e.g. “*Yes*”, “*No*”, or “*None*”) as well. For recognition of negated concepts (e.g. “patient denies having any fever”), KMCI implements NegEx, a widely-used algorithm to detect negations. However, NegEx may miss post-negation triggers such as “Cough: No”. We added regular expression rules based on our local note template to enhance negation detection.

The extended processing modules was implemented using Python 3.6. After processing, the extracted CUIs served as the input for following ConceptWAS analysis.

### ConceptWAS analysis

Similar like genome-wide association study (GWAS) and phenome-wide association study (PheWAS) that scan genomic and phenomic data for associations with a given disease or a genetic variant [16,17], conceptWAS examines the clinical concepts retrieved from clinical notes to determine if any concept is associated with a disease. In this study, we applied a conceptWAS to identify associations between symptoms-related concepts and the presence of COVID-19.

We applied Firth’s logistic regression to examine the association for each concept, adjusted by age, gender, and race. We chose Firth’s logistic regression because it has become a standard approach for analyzing binary outcomes with small samples [18]. Negated and non-negated CUIs are treated separately. A CUI with a negation flag (yes/no) is only counted once per patient. Firth’s logistic regression was implemented using R version 3.4.3 and the *logistf* package. As we tested multiple hypotheses, we used a Bonferroni correction for the significance level. We report the odds ratio (OR), p-values, and the prevalence in case and control groups for each CUI. We used a volcano plot to show p-values and the odds ratio for all CUIs. We also used a forest plot to show the significant concepts that were relevant to signs and symptoms.

### Chart review

We performed a manual chart review to evaluate the clinical plausibility of identified signals. We reviewed a CUI if 1) its p-value met Bonferroni-corrected significance, and 2) it was clinically meaningful (e.g., we excluded CUIs such as “*finding [CUI C00000243095]”* in a sentence like *“Findings are nonspecific*.*”*). We randomly selected notes from which the CUI was identified. Two authors (M.E.G. and H.N.E.) with clinical background ascertained whether the identified CUI was a true signal or false positive.

## Results

We identified 19,692 patients with COVID-19 PCR test results during the study period (Figure A.1). Patients’ mean age was 45 (44.6 ± 16.9) years. A total of 1,483 (7.5%) patients tested positive for COVID-19. The COVID-19-positive group was younger (41.5 ± 16.2 vs. 44.9 ± 16.9), more often male (48.0% vs. 41.7%), less often white (49.6% vs. 66.7%), and newer to VUMC (EHR length 7.3 years ± 8.1 vs. 9.2 ± 8.5) compared to COVID-19-negative patients (Table 1).

**Table 1.** Patient characteristics of the study cohort

| Attribute | Cases: COVID-19-positive<br>(n=1,483) | Controls: COVID-19-negative<br>(n=18,209) | P- value |
| --- | --- | --- | --- |
| Age (mean years +/- stddev) | 41.5 (16.2) | 44.9 (16.9) | <0.0001 |
| Gender (% Male) | 48.0% | 41.7% | <0.0001* |
| Race (% White) | 49.6% | 66.7% | <0.0001* |
| Average EHR length (years, +/- stddev) | 7.3 (8.1) | 9.2 (8.5) | <0.0001 |
| Average CUIs (+/- stddev) | 46.1 (61.1) | 71.9 (96.3) | <0.0001 |

\* 2-proportion z hypothesis test was performed. For age, EHR length, and average CUIs, a t- test was performed for comparing the median and standard deviations.

### Comparison of EHR-derived concepts between COVID-19 positive and negative patients

We extracted 87,753 clinical notes for the 19,692 patients. After using the NLP pipeline to processing the notes, we recognized 19,595 CUIs (including negated status) with semantic types of interests (Table B.1). Using ConceptWAS to compare EHR-derived concepts for COVID-19 positive and negative patients, 68 CUIs were identified after adjusting for multiple testing (Bonferroni-corrected significance, *P* < 2.55E-06) (Figure 1, Table E.1). The top signals included “depression” (OR = 0.34, 95% CI□=□0.24–0.47), “edema” (OR = 0.40, 95% CI = 0.29– 0.53), “fever (negated)” (OR = 0.63, 95% CI =□0.55–0.72), and “reaction anxiety” (OR = 0.39, 95% CI =□0.28–0.52). Specifically, symptoms concepts associated with COVID-19-positive patients included “absent sense of smell” (OR = 4.97, 95% CI = 3.21–7.50), “fever” (OR = 1.43, 95% CI = 1.28–1.59), “with cough fever” (OR = 2.29, 95% CI = 1.75–2.96), and “ageusia” (OR = 5.18, 95% CI = 3.02–8.58) (Figure 2).

**Figure 1.**
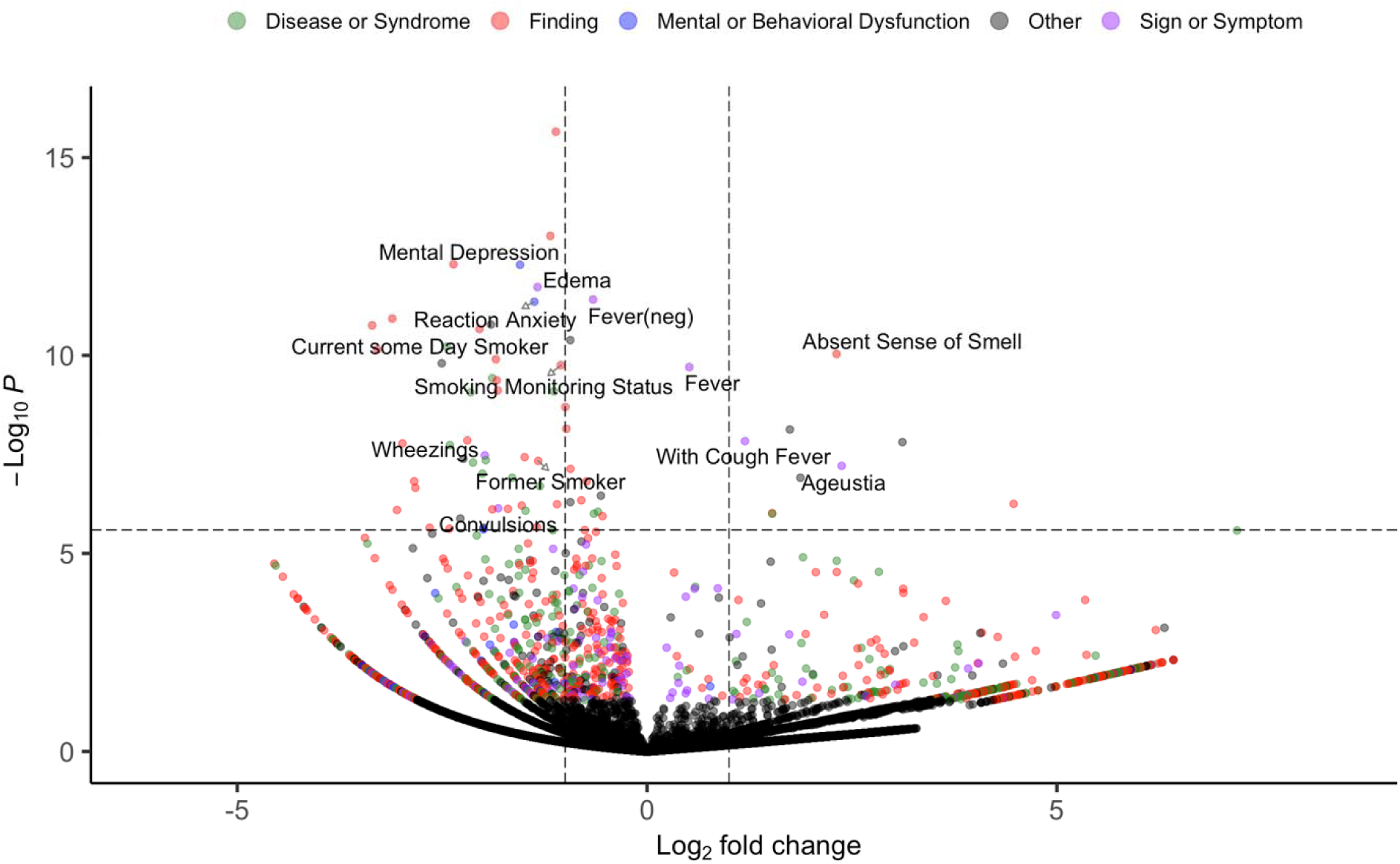
Volcano plot of a ConceptWAS scan for 19, 692 patients that included COVID-19-positive group (cases) and negative group (controls). The points are colored by the semantic type of the concepts. Selected associations related to signs, symptoms, or diseases/syndromes are labeled. The volcano plot indicates -log 10 (p-value) for association (y-axis) plotted against their respective log 2 (fold change) (x-axis). The dashed line represents significance level using a Bonferroni correction.

**Figure 2.**
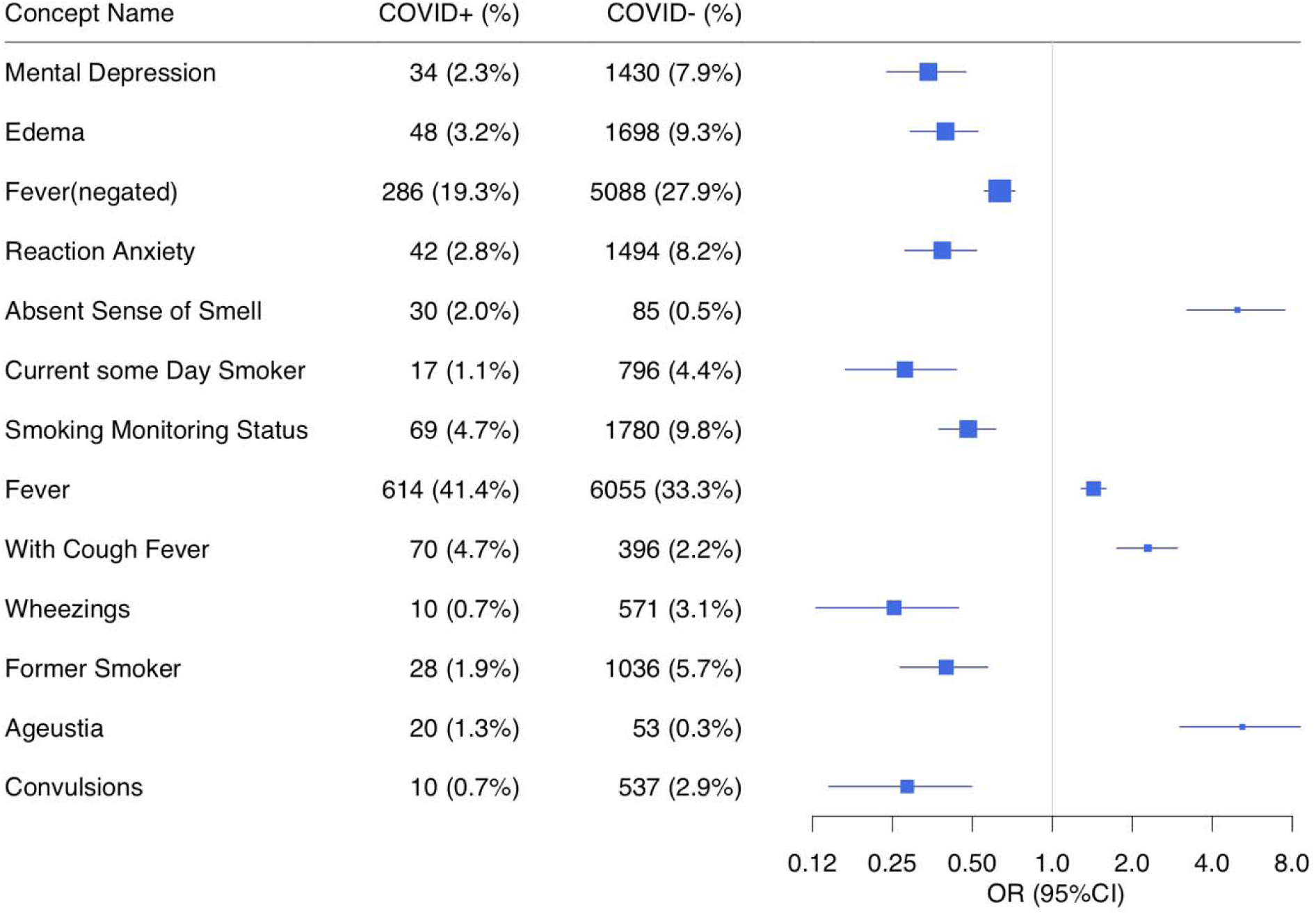
Forest plot comparing individual concepts between COVID-19-positive (case) and COVID-19-negative (control) patients. Selected associations include the significant signals related to semantic types of symptoms that met Bonferroni-corrected significance (p-value < 2.55E-06). The odds ratio has been adjusted for age, gender, and race. The concepts are ordered by p-value.

Concepts related to smoking status such as “current some day smoker”, “former smoker”, and “smoking monitoring status” were more frequently reported in the COVID-negative group than in the COVID-positive group (OR < 1, *P* < 2.55E-06), suggesting more smokers in control group. To ascertain whether this signal was true or false positives due to wrongly assertion detection by NLP pipeline, we performed a chart review of 80 patients’ notes that had smoking-related CUIs. We found that 79 of 80 patients confirmed an affirmative smoking status (see below chart review).

### Temporal analysis

We performed ConceptWAS using the cumulative data every two weeks within the study period (Figure 3, Figure D.1). By week 4 (by April 5, 2020), “absence of smell” (OR□=□10.24; 95% CI□=□5.18–20.06) and “ageusia” (loss of taste, OR =□11.79; 95% CI =□5.55–25.2) became significantly associated with increased risk of COVID-19 infection. These two signals remained significant through the subsequent weeks (Supplementary Data). Fever (negated) appeared (OR =□0.55; 95% CI =□0.43–0.71) at week 2 (between March 8 and 22, 2020), and “with cough fever” became significant (OR =□2.09; 95% CI =□1.60–2.70) from the 8th week (between March 8 and May 3, 2020). The depression and anxiety appeared significantly starting from week 4 (by April 5, 2020).

**Figure 3.**
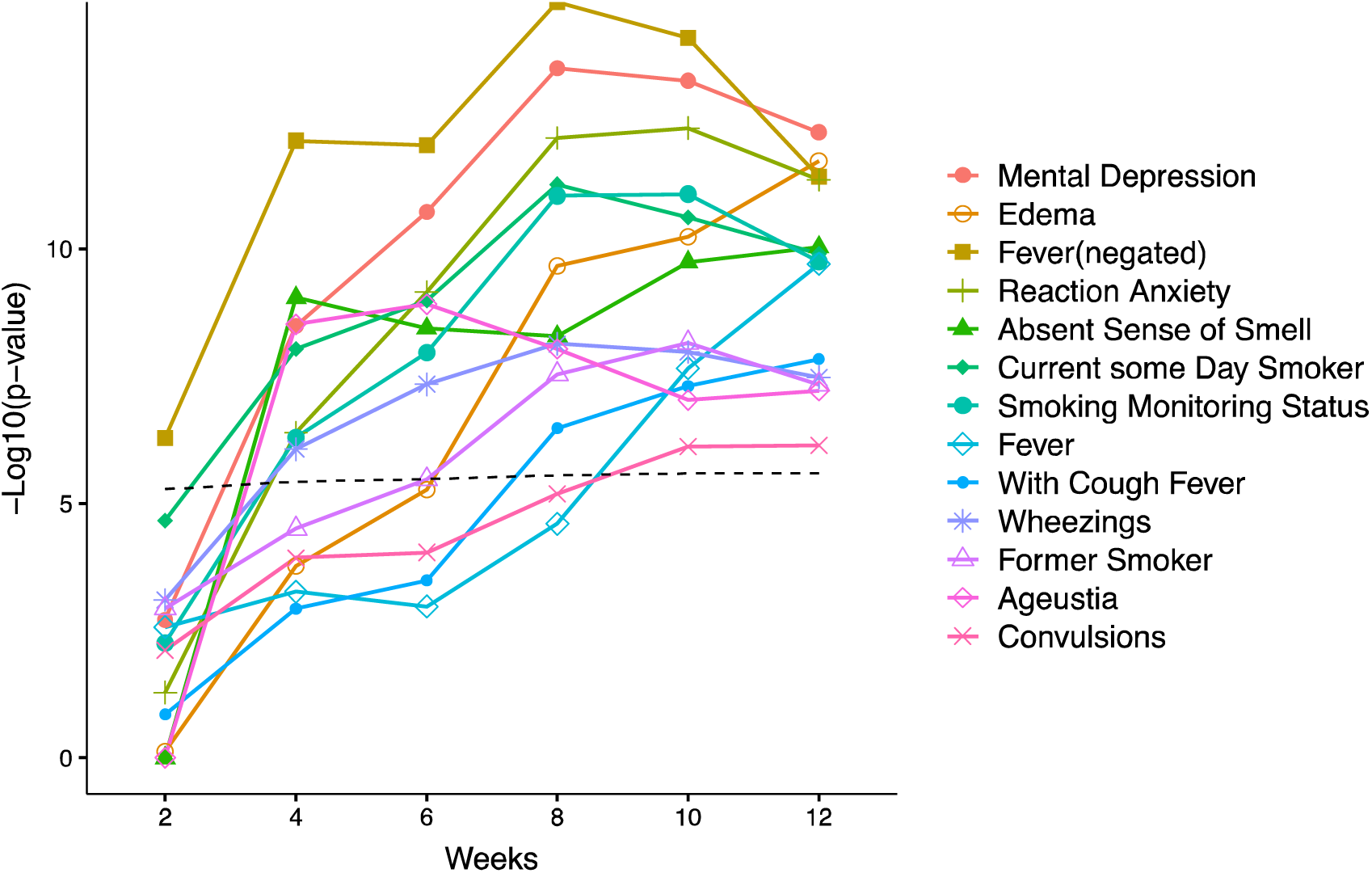
Temporal ConceptWAS using bi-weekly cumulative data. For significant signals (related to signs, symptoms) using all data (labeled in Figure 2), the plot indicates their -log 10 (p-value) for association (y-axis) against using the cumulative data started between March 8, 2020 to *n* weeks (x-axis). The dashed line indicates a significant association using a Bonferroni correction.

### Chart review

To validate the signals, we reviewed patient’s charts for significant concepts with high clinical relevance. We randomly selected 20 notes for each concept to review whether the notes mentioned the symptoms in the expected attribute (e.g. affirmative or negated) (Table 2). The significant concepts from the ConceptWAS that compared COVID-19 positive and negative patients, such as “absent sense of smell”, “ageusia”, “mental depression”, and concepts related to smoking status (e.g. “current some day smoker”, “former smoker”, and “smoking monitoring status”) were consistent with the expected attribute based on chart review.

**Table 2.** Results of chart reviews.

| Concepts | Reviewed samples | True signals | True signals percentage % | Examples of false positive |
| --- | --- | --- | --- | --- |
| Absent sense of smell | 20 | 19 | 95.00% | "(-) altered/loss of smell", were wrongly recognized as an affirmative/ positive attribute. |
| Ageusia | 20 | 19 | 95.00% | "Symptoms, n/v, fever, cough, loss of taste or smell or around anyone + for Covid 19." |
| Mental Depression | 20 | 18 | 90.00% | One was recognized from a medical history title without any answers; the other came from a recommendation for further Psychosocial assessment. |
| Current some day smoker | 20 | 20 | 100.00% |  |
| Smoking monitoring status | 20 | 19 | 95.00% | One is uncertain. "Smoking Status Not on file" . |
| Fever | 20 | 17 | 85.00% | Template issue. "The following ROS were reviewed and are negative, unless otherwise stated as +positive:<br>1 Constitutional: Fever; malaise" |
| Pericardial Fluid (neg) | 20 | 20 | 100.00% |  |
| Hydrocephalus [neg] | 20 | 20 | 100.00% |  |
| Hydronephroses | 20 | 20 | 100.00% |  |
| Blood group AB Rh(D) negative (finding) | 20 | 0 | 0.00% | From blood typing tests. This signal was not specific to blood type AB+, but generated by other ABO blood types and Rh-positive patients. |
| Allergy test positive (finding) | 20 | 5 | 25.00% | The false positives were wrongly mapped from a sentence like "He /She has been exposed to covid, family member or friends have tested positive." |
| Laurin-Sandrow syndrome | 20 | 20 | 100.00% |  |
| Cough nonproductive | 20 | 20 | 100.00% |  |
| In total | 260 | 217 | 83.46% |  |

Although “smoking monitoring status” was generated by an inquiry term used in a template of a chart, after we post-processed the KMCI output to remove irrelevant concepts and refine negation, the smoking monitoring status followed by a negated answer was recognized as a negated attribute. We reviewed 20 notes that mentioned the “smoking monitoring status (affirmative/positive attribute)” and 19 were either current or former smokers.

We also found false positive concepts, mostly due to NLP entity recognition errors. For example, “additional information” was recognized as “adequate knowledge”. The concept “fever” with positive attribute has three false positives, mainly due to a few specific chart templates used for denoting the negation, which were not captured by NLP pipeline.

## Discussion

Our work describes a high-throughput and reproducible approach (ConceptWAS) that use EHR notes to early identify pandemic disease symptoms and investigate clinical manifestations for further hypothesis-driven study. We applied ConceptWAS to a cohort of patients who underwent COVID-19 testing. We replicated several well-known symptoms of COVID-19, such as fever, loss of smell/taste, and cough with fever [19–21]. Using ConceptWAS, we were able to detect the signal of loss of smell and taste as early as April 5, 2020, nearly three weeks earlier than the date that they were listed as COVID-19 symptoms by the CDC [3]. - Our results demonstrate the feasibility of using ConceptWAS for an early detection of symptoms of an unknown disease.

We also observed several signals enriched in the COVID-19-negative group. For example, depression and anxiety have a higher prevalence among patients who tested negative. These signals first became significant starting from April 5, 2020, corresponding to the date when the Governor of Tennessee issued a “safer at home” Executive Order and a “stay at home” order. It reflects the mental health issues that the shutdown and quarantine policies may bring to the people [22,23]. We also find a higher percentage of smoking status concepts in the COVID-19-negative group. Earlier epidemiological studies found that fewer smokers are among COVID-19 patients or hospitalized COVID-19 patients [21,24], which are consistent with our findings of the negative correlation between smoking and COVID-19. One explanation could be the impact of nicotine on ACE-2, as nicotine has been suggested to play a protective role against COVID-19 [25]. It is also possible that smokers are taking greater social precautions because of perceived higher risk for respiratory complications from COVID-19, thus reducing their risk of contracting the virus. Although these findings suggest that smoking may be a protective factor, lack of evidence and known adverse events associated with smoking dissuade continued smoking as a protective measure against COVID-19.

ConceptWAS is open-source, portable, and reproducible. Researchers/Users can choose other NLP pipelines (e.g. MetaMap, CLAMP, cTAKES) [26] for concept extraction and use the derived concepts as the input to ConceptWAS.

Through a proof-of-concept study by applying NLP techniques to identify COVID-19 symptoms, we summarize the lessons learned that may help others apply this method.

1. A high-throughput, lightweight, and reproducible method is important for pandemic disease. ConceptWAS enables a rapid scan of symptoms using clinical notes. These symptoms provided an initial hypothesis for further investigation and could alert clinicians to pay attention to patients with presenting specific symptoms. Researchers can run the ConceptWAS regularly (e.g., weekly or bi-weekly cumulative data) to track the symptoms changes for a pandemic disease
2. Running ConceptWAS needs to be cautious about the distribution of different clinical note types. Different clinical note types are unlike from each other due to their specific clinical usage. They may have variable templates and inconsistent length. Therefore, we recommend that researchers check the distribution of document types between cases and controls to avoid sampling bias
3. Although NLP has been used in various medical fields to improve information processing and practice [26–29], recognition of negative and uncertain concepts remains a challenge. We enhanced the detection of uncertain arguments and negated concepts by developing rule-based methods as wrappers for entity-identification generated results. Still, our manual chart review suggest that the outcome is not perfect. For example, some notes mentioned negative concepts like “the following ROS were reviewed and are negative, unless otherwise stated as +positive: Constitutional: Fever; malaise.” Such scenarios are difficult for NLP tools to identify. A combination of machine learning and rule-based approaches may improve the detection
4. We also learned and recognized that our study has several limitations. First, the study was performed at a single institution with a limited number of COVID-19 patients. As the pandemic crisis evolves and more patients are tested for SARS-CoV-2 in our healthcare system, our ability to detect COVID-19 and clinical concepts’ associations will continue to improve. Second, this study used data from a limited time (before May 27, 2020). In the future, we will extract notes prior to the test date to study the progression of the symptoms and analyze them along different periods. Lastly, as the performance of an NLP system may vary across institutions and databases [26,30], further studies are necessary to assess the generalizability of our findings

## Conclusion

In this study, we describe a high-throughput approach (ConceptWAS) that systematically scans a disease’s clinical manifestations from clinical notes. By applying ConceptWAS on EHR clinical notes from patients subjecting to a COVID-19 test, we detected loss of smell/taste three weeks prior to their inclusion as symptoms of the disease by CDC. The study demonstrates the potential of the EHR-based methods to enable early recognition of specific COVID-19 symptoms, and improve our clinical and public health response to the pandemic.

## Data Availability

The study used data from patients represented in the Vanderbilt University Medical Center.The study was approved by the Vanderbilt University Medical Center Institutional Review Board (IRB #200512). The summary statistics derived from the EHRs are enclosed within the manuscript.

## Code availability

Up-to-date developments of ConceptWAS are available in GitHub (https://github.com/zhaojuanwendy/ConceptWAS).

## Acknowledgments

We thanked Dr. Vivian Siegel from Department of Biology at MIT Department of Medicine at Vanderbilt University for helpful suggestions on the study design and manuscript drafting.

## Funding

The project was supported by the National Institutes of Health (NIH), National Institute of General Medical Studies (P50 GM115305), National Heart, Lung, and Blood Institute (R01 HL133786), National Library of Medicine (T15 LM007450, R01 LM010685), and the American Heart Association (18AMTG34280063), as well as the Vanderbilt Biomedical Informatics Training Program, Vanderbilt Faculty Research Scholar Fund, and the Vanderbilt Medical Scientist Training Program. The datasets used for the analyses described were obtained from Vanderbilt University Medical Center’s resources and the Synthetic Derivative, which are supported by institutional funding and by the Vanderbilt National Center for Advancing Translational Science grant (UL1 TR000445) from NCATS/NIH. The funders had no role in study design, data collection and analysis, decision to publish, or preparation of the manuscript.

## Contributions

J.Z. and W.Q.W. developed the ConceptWAS concept and methodology. J.Z. and M.E.G. wrote the code, performed analysis, and plotted the figure. J.Z. and J.C.S. developed the ConceptWAS NLP pipelines. V.E.K. and W.Q.W. interpreted the clinical meaning of the results. M.E.G. and H.N.E. performed the chart review. V.E.K, Q.P., J.P., S.T.R. and K.B.J. provided the feedback. J.Z., M.E.G., V.E.K., J.C.S., H.N.E., Q.P., J.P., S.T.R., K.B.J., and W.Q.W. wrote and edited the manuscript.

## Competing interests

The authors have no competing interests to declare.

## Notes

### Competing Interest Statement

The authors have declared no competing interest.

### Author Declarations

RE: IRB #200731 "EHR Patterns of COVID-19" Dear Wei-qi Wei: A designee of the Institutional Review Board reviewed the Request for Exemption application identified above. It was determined the study poses minimal risk to participants. This study meets 45 CFR 46.104 (d) category (4) for Exempt Review. Any changes to this proposal that may alter its exempt status should be presented to the IRB for approval prior to implementation of the changes. DATE OF IRB APPROVAL: 5/21/2020 Sincerely, Erin Leigh Johnson BS, CIP Institutional Review Board Behavioral Sciences Committee Electronic Signature: Erin Leigh Johnson/VUMC/Vanderbilt : (2c8072da56409aeae2a38cff74965eed) Signed On: 05/21/2020 3:24:30 PM CDT

## References

[1] WHO Coronavirus Disease (COVID-19) Dashboard, (n.d.). https://covid19.who.int/ (accessed May 26, 2020).

[2] W. Guan, Z. Ni, Y. Hu, W. Liang, C. Ou, J. He, L. Liu, H. Shan, C. Lei, D.S.C. Hui, B. Du, L. Li, G. Zeng, K.-Y. Yuen, R. Chen, C. Tang, T. Wang, P. Chen, J. Xiang, S. Li, J. Wang, Z. Liang, Y. Peng, L. Wei, Y. Liu, Y. Hu, P. Peng, J. Wang, J. Liu, Z. Chen, G. Li, Z. Zheng, S. Qiu, J. Luo, C. Ye, S. Zhu, N. Zhong, Clinical Characteristics of Coronavirus Disease 2019 in China, New England Journal of Medicine. (2020). https://doi.org/10.1056/NEJMoa2002032.

[3] J. Makaronidis, J. Mok, N. Balogun, C.G. Magee, R.Z. Omar, A. Carnemolla, R.L. Batterham, Seroprevalence of SARS-CoV-2 antibodies in people with an acute loss in their sense of smell and/or taste in a community-based population in London, UK: An observational cohort study, PLOS Medicine. 17 (2020) e1003358. https://doi.org/10.1371/journal.pmed.1003358.

[4] A. Fritz, M. Brice-Saddler, M. Judkis, CDC confirms six coronavirus symptoms showing up in patients over and over, Washington Post. (n.d.). https://www.washingtonpost.com/health/2020/04/27/six-new-coronavirus-symptoms/ (accessed September 25, 2020).

[5] Statement from the UK Chief Medical Officers on an update to coronavirus symptoms: 18 May 2020, GOV.UK. (n.d.). https://www.gov.uk/government/news/statement-from-the-uk-chief-medical-officers-on-an-update-to-coronavirus-symptoms-18-may-2020 (accessed June 5, 2020).

[6] R. Awasthi, R. Pal, P. Singh, A. Nagori, S. Reddy, A. Gulati, P. Kumaraguru, T. Sethi, CovidNLP: A Web Application for Distilling Systemic Implications of COVID-19 Pandemic with Natural Language Processing, MedRxiv. (2020) 2020.04.25.20079129. https://doi.org/10.1101/2020.04.25.20079129.

[7] T. Mackey, V. Purushothaman, J. Li, N. Shah, M. Nali, C. Bardier, B. Liang, M. Cai, R. Cuomo, Machine Learning to Detect Self-Reporting of Symptoms, Testing Access, and Recovery Associated With COVID-19 on Twitter: Retrospective Big Data Infoveillance Study, JMIR Public Health Surveill. 6 (2020). https://doi.org/10.2196/19509.

[8] R.M. Burke, Symptom Profiles of a Convenience Sample of Patients with COVID-19 — United States, January–April 2020, MMWR Morb Mortal Wkly Rep. 69 (2020). https://doi.org/10.15585/mmwr.mm6928a2.

[9] C. Menni, A.M. Valdes, M.B. Freidin, C.H. Sudre, L.H. Nguyen, D.A. Drew, S. Ganesh, T. Varsavsky, M.J. Cardoso, J.S. El-Sayed Moustafa, A. Visconti, P. Hysi, R.C.E. Bowyer, M. Mangino, M. Falchi, J. Wolf, S. Ourselin, A.T. Chan, C.J. Steves, T.D. Spector, Real-time tracking of self-reported symptoms to predict potential COVID-19, Nature Medicine. 26 (2020) 1037–1040. https://doi.org/10.1038/s41591-020-0916-2.

[10] S. Richardson, J.S. Hirsch, M. Narasimhan, J.M. Crawford, T. McGinn, K.W. Davidson, D.P. Barnaby, L.B. Becker, J.D. Chelico, S.L. Cohen, J. Cookingham, K. Coppa, M.A. Diefenbach, A.J. Dominello, J. Duer-Hefele, L. Falzon, J. Gitlin, N. Hajizadeh, T.G. Harvin, D.A. Hirschwerk, E.J. Kim, Z.M. Kozel, L.M. Marrast, J.N. Mogavero, G.A. Osorio, M. Qiu, T.P. Zanos, Presenting Characteristics, Comorbidities, and Outcomes Among 5700 Patients Hospitalized With COVID-19 in the New York City Area, JAMA. 323 (2020) 2052– 2059. https://doi.org/10.1001/jama.2020.6775.

[11] G.A. Brat, G.M. Weber, N. Gehlenborg, P. Avillach, N.P. Palmer, L. Chiovato, J. Cimino, L.R. Waitman, G.S. Omenn, A. Malovini, J.H. Moore, B.K. Beaulieu-Jones, V. Tibollo, S.N. Murphy, S. L’Yi, M.S. Keller, R. Bellazzi, D.A. Hanauer, A. Serret-Larmande, A. Gutierrez-Sacristan, J.H. Holmes, D.S. Bell, K.D. Mandl, R.W. Follett, J.G. Klann, D.A. Murad, L. Scudeller, M. Bucalo, K. Kirchoff, J. Craig, J. Obeid, V. Jouhet, R. Griffier, S. Cossin, B. Moal, L.P. Patel, A. Bellasi, H.U. Prokosch, D. Kraska, P. Sliz, A.L. Tan, K.Y. Ngiam, A. Zambelli, D.L. Mowery, E. Schiver, B. Devkota, R.L. Bradford, M. Daniar, APHP/Universities/INSERM COVID-19 research collaboration, C. Daniel, V. Benoit, R. Bey, N. Paris, A.S. Jannot, P. Serre, N. Orlova, J. Dubiel, M. Hilka, A.S. Jannot, S. Breant, J. Leblanc, N. Griffon, A. Burgun, M. Bernaux, A. Sandrin, E. Salamanca, T. Ganslandt, T. Gradinger, J. Champ, M. Boeker, P. Martel, A. Gramfort, O. Grisel, D. Leprovost, T. Moreau, G. Varoquaux, J.-J. Vie, D. Wassermann, A. Mensch, C. Caucheteux, C. Haverkamp, G. Lemaitre, I.D. Krantz, S. Cormont, A. South, The Consortium for Clinical Characterization of COVID-19 by EHR (4CE), T. Cai, I.S. Kohane, International Electronic Health Record-Derived COVID-19 Clinical Course Profiles: The 4CE Consortium, Infectious Diseases (except HIV/AIDS), 2020. https://doi.org/10.1101/2020.04.13.20059691.

[12] T. Wagner, F. Shweta, K. Murugadoss, S. Awasthi, A. Venkatakrishnan, S. Bade, A. Puranik, M. Kang, B.W. Pickering, J.C. O’Horo, P.R. Bauer, R.R. Razonable, P. Vergidis, Z. Temesgen, S. Rizza, M. Mahmood, W.R. Wilson, D. Challener, P. Anand, M. Liebers, Z. Doctor, E. Silvert, H. Solomon, A. Anand, R. Barve, G. Gores, A.W. Williams, W.G. Morice II, J. Halamka, A. Badley, V. Soundararajan, Augmented curation of clinical notes from a massive EHR system reveals symptoms of impending COVID-19 diagnosis, ELife. 9 (2020) e58227. https://doi.org/10.7554/eLife.58227.

[13] J.C. Denny, A. Spickard, R.A. Miller, J. Schildcrout, D. Darbar, S.T. Rosenbloom, J.F. Peterson, Identifying UMLS concepts from ECG Impressions using KnowledgeMap, AMIA Annu Symp Proc. (2005) 196–200.

[14] J.C. Denny, P.R. Irani, F.H. Wehbe, J.D. Smithers, A. Spickard, The KnowledgeMap Project: Development of a Concept-Based Medical School Curriculum Database, AMIA Annu Symp Proc. 2003 (2003) 195–199.

[15] J.C. Denny, J.F. Peterson, N.N. Choma, H. Xu, R.A. Miller, L. Bastarache, N.B. Peterson, Extracting timing and status descriptors for colonoscopy testing from electronic medical records, J Am Med Inform Assoc. 17 (2010) 383–388. https://doi.org/10.1136/jamia.2010.004804.

[16] L.A. Hindorff, P. Sethupathy, H.A. Junkins, E.M. Ramos, J.P. Mehta, F.S. Collins, T.A. Manolio, Potential etiologic and functional implications of genome-wide association loci for human diseases and traits, PNAS. 106 (2009) 9362–9367. https://doi.org/10.1073/pnas.0903103106.

[17] J.C. Denny, M.D. Ritchie, M.A. Basford, J.M. Pulley, L. Bastarache, K. Brown-Gentry, D. Wang, D.R. Masys, D.M. Roden, D.C. Crawford, PheWAS: demonstrating the feasibility of a phenome-wide scan to discover gene–disease associations, Bioinformatics. 26 (2010) 1205–1210. https://doi.org/10.1093/bioinformatics/btq126.

[18] Firth’s logistic regression with rare events: accurate effect estimates and predictions? – Puhr - 2017 - Statistics in Medicine - Wiley Online Library, (n.d.). https://onlinelibrary.wiley.com/doi/full/10.1002/sim.7273 (accessed June 7, 2020).

[19] COVID-19 Patients’ Clinical Characteristics, Discharge Rate, and Fatality Rate of Meta-Analysis - PubMed, (n.d.). https://pubmed.ncbi.nlm.nih.gov/32162702/ (accessed June 30, 2020).

[20] L.A. Vaira, G. Salzano, G. Deiana, G. De Riu, Anosmia and Ageusia: Common Findings in COVID-19 Patients, Laryngoscope. 130 (2020) 1787. https://doi.org/10.1002/lary.28692.

[21] S.T. Moein, S.M. Hashemian, B. Mansourafshar, A. Khorram-Tousi, P. Tabarsi, R.L. Doty, Smell dysfunction: a biomarker for COVID-19, International Forum of Allergy & Rhinology. n/a (n.d.). https://doi.org/10.1002/alr.22587.

[22] B. Pfefferbaum, NC.S. orth, Mental Health and the Covid-19 Pandemic, New England Journal of Medicine. 383 (2020) 510–512. https://doi.org/10.1056/NEJMp2008017.

[23] W. Sturges, Gov. Bill Lee issues stay-at-home order through April 14, Impact. (2020). https://communityimpact.com/nashville/franklin-brentwood/coronavirus/2020/03/30/gov-bill-lee-issues-statewide-stay-at-home-order-for-tennesseans/ (accessed October 7, 2020).

[24] A. Emami, F. Javanmardi, N. Pirbonyeh, A. Akbari, Prevalence of Underlying Diseases in Hospitalized Patients with COVID-19: a Systematic Review and Meta-Analysis, Arch Acad Emerg Med. 8 (2020). https://www.ncbi.nlm.nih.gov/pmc/articles/PMC7096724/ (accessed July 31, 2020).

[25] K. Farsalinos, R. Niaura, J. Le Houezec, A. Barbouni, A. Tsatsakis, D. Kouretas, A. Vantarakis, K. Poulas, Editorial: Nicotine and SARS-CoV-2: COVID-19 may be a disease of the nicotinic cholinergic system, Toxicol Rep. 7 (2020) 658–663. https://doi.org/10.1016/j.toxrep.2020.04.012.

[26] E. Soysal, J. Wang, M. Jiang, Y. Wu, S. Pakhomov, H. Liu, H. Xu, CLAMP - a toolkit for efficiently building customized clinical natural language processing pipelines, J Am Med Inform Assoc. 25 (2018) 331–336. https://doi.org/10.1093/jamia/ocx132.

[27] M. Yetisgen-Yildiz, M.L. Gunn, F. Xia, T.H. Payne, Automatic identification of critical follow-up recommendation sentences in radiology reports, AMIA Annu Symp Proc. 2011 (2011) 1593–1602.

[28] D. Mf, S. S, B. Ws, D. Jc, H. Jl, Automated extraction of clinical traits of multiple sclerosis in electronic medical records, Journal of the American Medical Informatics Association□: JAMIA. 20 (2013). https://doi.org/10.1136/amiajnl-2013-001999.

[29] Y. Ww, Y. M, H. Wp, K. Sw, Natural Language Processing in Oncology: A Review, JAMA Oncology. 2 (2016). https://doi.org/10.1001/jamaoncol.2016.0213.

[30] Negation’s Not Solved: Generalizability Versus Optimizability in Clinical Natural Language Processing, (n.d.). https://journals.plos.org/plosone/article?id=10.1371/journal.pone.0112774 (accessed August 18, 2020).

