## Supplementary material for "ConceptWAS: a high-throughput method for early identification of COVID-19 presenting symptoms": Figure A.1;Figure B.1;Table C.1;Figure D.1;Table E.1

**Appendix A. Study design**

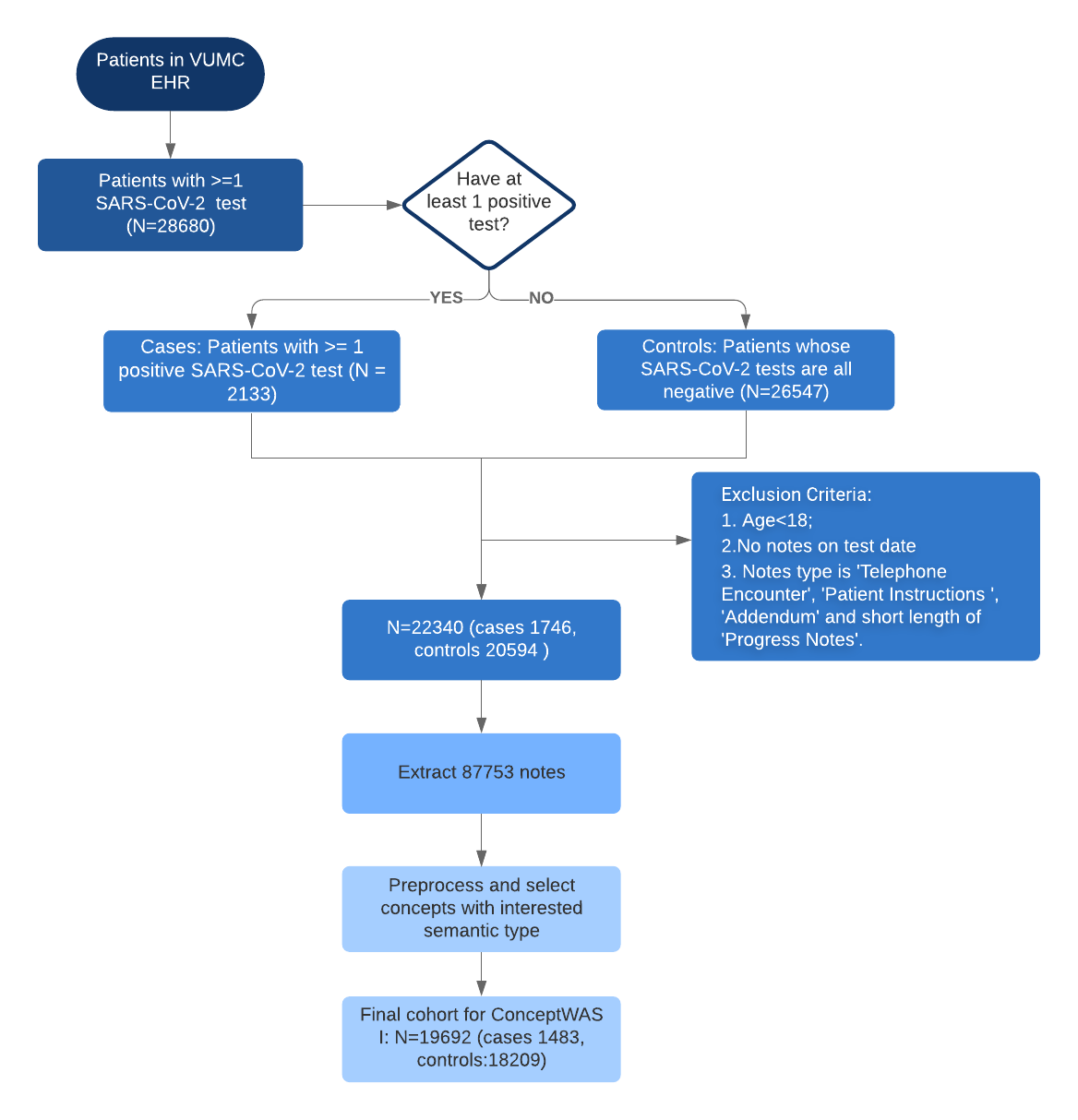
<https://app.lucidchart.com/documents/edit/b4f34f20-62ec-41b0-a291-ea050bd8ecd6/0?callback=close&name=docs&callback_type=back&v=1651&s=584>

**Figure A.1.** Flowchart of study design for ConceptWAS between COVID-19-positive (case) and COVID-19 negatives (control).

**Appendix B. EHR notes distribution**

The COVID-19-positive group had a distribution of clinical notes types similar to that of the COVID-19-negative group on the PCR test day (e.g. progress notes 81.36% versus [vs] 73.73%, social history 22.95% vs 28.41%, emergency department [ED] provider notes 6.26% vs 7.98%).

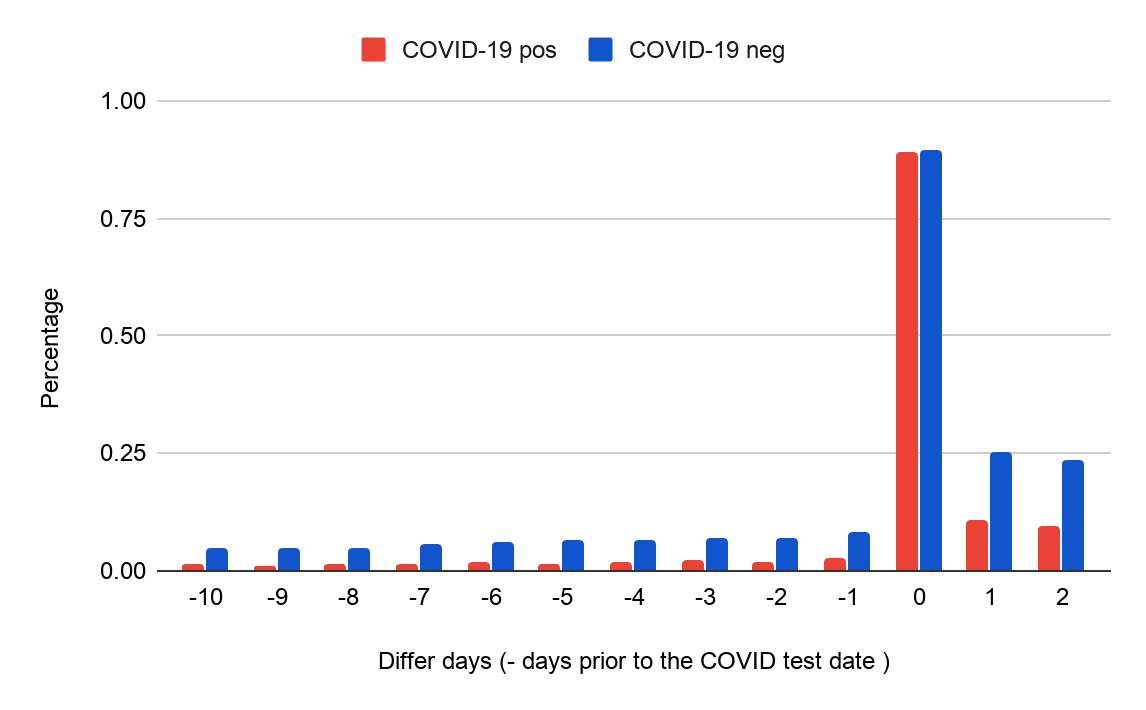

**Figure B.1.** Proportion of cases/controls with clinical notes on the days around COVID-19 test date. The x-axis indicates the note day relative to the COVID-19 test date. >86% patients who have a PCR test had a clinical note within 24 hours before the test date.

**Appendix C. Framework of the NLP pipeline and ConceptWAS**

**Figure C.1.** Schematic framework of the ConceptWAS

**Table C.1.** Semantic type of concepts that were included in the analysis.

**Semantic type**

Sign or Symptom

Finding

Disease or Syndrome

Mental Process

Mental or Behavioral Dysfunction

Organism Function

Laboratory or Test Result

Individual Behavior

Social Behavior

Acquired Abnormality

Age Group

Population Group

**Appendix D Temporal analysis**

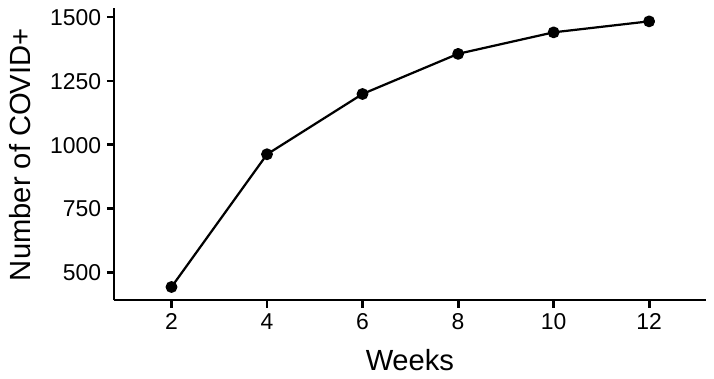

**Figure D.1.** The cumulative number of COVID-19-positive(cases) along weeks.

**Appendix E. ConceptWAS results**

**Table E.1.** ConceptWAS between COVID-19-positive (case) and COVID-19 negative(control). The table presents the significant concepts related to sign or symptom, disease or syndrome, or individual behaviours, which crossed Bonferroni p-value < 2.55E-06. It also includes concepts with semantic type of symptoms, which were enriched in COVID-19 and p-value<0.05.

| Concept Name | Semantic type | Case Count (%) | Control Count(%) | OR (95%CI) | P-value |
| --- | --- | --- | --- | --- | --- |
| Adequate Knowledge | Finding | 109 (7.3%) | 3033 (16.7%) | 0.46 (0.38,0.56) | 2.22E-16 |
| Finding | Finding | 75 (5.1%) | 2309 (12.7%) | 0.44 (0.34,0.56) | 9.55E-14 |
| Increased thickness (Finding) | Finding | 11 (0.7%) | 854 (4.7%) | 0.19 (0.10,0.33) | 4.97E-13 |
| Mental depression | Mental or Behavioral Dysfunction | 34 (2.3%) | 1430 (7.9%) | 0.34 (0.24,0.47) | 5.12E-13 |
| Edema | Sign or Symptom | 48 (3.2%) | 1698 (9.3%) | 0.40 (0.29,0.53) | 1.88E-12 |
| Fever(negated) | Sign or Symptom | 286 (19.3%) | 5088 (27.9%) | 0.63 (0.55,0.72) | 3.87E-12 |
| Reaction anxiety | Mental or Behavioral Dysfunction | 42 (2.8%) | 1494 (8.2%) | 0.39 (0.28,0.52) | 4.40E-12 |
| Anesthesia Type (Navigational Concept) | Finding | 4 (0.3%) | 536 (2.9%) | 0.12 (0.04,0.26) | 1.17E-11 |
| Result, Lab.- General (Observable Entity) | Laboratory or Test Result | 17 (1.1%) | 891 (4.9%) | 0.27 (0.16,0.42) | 1.65E-11 |
| Neck Flexion (Finding) | Finding | 3 (0.2%) | 494 (2.7%) | 0.10 (0.03,0.24) | 1.74E-11 |
| Crisis | Finding | 14 (0.9%) | 850 (4.7%) | 0.24 (0.14,0.39) | 2.16E-11 |
| Finding of tobacco smoking behavior (Finding) | Individual Behavior | 99 (6.7%) | 2445 (13.4%) | 0.52 (0.42,0.64) | 4.11E-11 |
| Hydrocephalus [disease/Finding](negated) | Disease or Syndrome | 8 (0.5%) | 662 (3.6%) | 0.18 (0.09,0.34) | 5.98E-11 |
| Sedated State(negated) | Finding | 3 (0.2%) | 445 (2.4%) | 0.10 (0.03,0.25) | 7.07E-11 |
| Absent sense of smell | Finding | 30 (2.0%) | 85 (0.5%) | 4.97 (3.21,7.50) | 9.21E-11 |
| Current some day smoker | Finding | 17 (1.1%) | 796 (4.4%) | 0.28 (0.17,0.44) | 1.25E-10 |
| Blood Group Ab Rh(d) Negative (Finding) | Laboratory or Test Result | 7 (0.5%) | 589 (3.2%) | 0.18 (0.08,0.34) | 1.58E-10 |
| Smoking monitoring status | Finding | 69 (4.7%) | 1780 (9.8%) | 0.48 (0.37,0.61) | 1.77E-10 |
| Fever | Sign or Symptom | 614 (41.4%) | 6055 (33.3%) | 1.43 (1.28,1.59) | 1.97E-10 |
| Pericardial Fluid(negated) | Disease or Syndrome | 15 (1.0%) | 819 (4.5%) | 0.27 (0.16,0.43) | 3.72E-10 |
| Postures | Finding | 16 (1.1%) | 821 (4.5%) | 0.28 (0.16,0.44) | 4.26E-10 |
| Lesion | Finding | 16 (1.1%) | 860 (4.7%) | 0.28 (0.17,0.45) | 7.68E-10 |
| Consolidation of Lung(negated) | Disease or Syndrome | 53 (3.6%) | 1604 (8.8%) | 0.46 (0.34,0.60) | 8.03E-10 |
| Hydronephroses(negated) | Disease or Syndrome | 10 (0.7%) | 667 (3.7%) | 0.22 (0.11,0.39) | 8.57E-10 |
| White Blood Cell Count Increased (Lab Result) | Finding | 70 (4.7%) | 1951 (10.7%) | 0.50 (0.39,0.64) | 2.02E-09 |
| Comments on own reading (Finding) | Finding | 66 (4.5%) | 1850 (10.2%) | 0.51 (0.39,0.65) | 7.13E-09 |
| Allergy test positive (Finding) | Laboratory or Test Result | 39 (2.6%) | 128 (0.7%) | 3.35 (2.29,4.79) | 7.42E-09 |
| Disability | Finding | 8 (0.5%) | 559 (3.1%) | 0.22 (0.10,0.40) | 1.40E-08 |
| With cough fever | Sign or Symptom | 70 (4.7%) | 396 (2.2%) | 2.29 (1.75,2.96) | 1.46E-08 |
| Sickling test positive (Finding) | Laboratory or Test Result | 15 (1.0%) | 22 (0.1%) | 8.66 (4.38,16.69) | 1.55E-08 |
| Patient condition unchanged (Finding) | Finding | 3 (0.2%) | 403 (2.2%) | 0.13 (0.04,0.31) | 1.67E-08 |
| Paresthesias(negated) | Disease or Syndrome | 6 (0.4%) | 441 (2.4%) | 0.19 (0.08,0.38) | 1.84E-08 |
| Wheezings | Sign or Symptom | 10 (0.7%) | 571 (3.1%) | 0.25 (0.13,0.44) | 3.35E-08 |
| Does with Much Difficulty (Qualifier Value) | Finding | 21 (1.4%) | 871 (4.8%) | 0.35 (0.22,0.53) | 3.71E-08 |
| Nodule | Acquired Abnormality | 7 (0.5%) | 552 (3.0%) | 0.21 (0.09,0.41) | 4.08E-08 |
| Leukocytosis | Disease or Syndrome | 10 (0.7%) | 607 (3.3%) | 0.26 (0.13,0.45) | 4.42E-08 |
| Former smoker | Finding | 28 (1.9%) | 1036 (5.7%) | 0.40 (0.27,0.57) | 4.62E-08 |
| Epileptic seizures | Disease or Syndrome | 8 (0.5%) | 556 (3.1%) | 0.23 (0.11,0.42) | 5.09E-08 |
| Ageustia | Sign or Symptom | 20 (1.3%) | 53 (0.3%) | 5.18 (3.02,8.58) | 6.16E-08 |
| both patent | Finding | 64 (4.3%) | 1673 (9.2%) | 0.52 (0.40,0.67) | 7.36E-08 |
| Pleural effusion disorder | Disease or Syndrome | 9 (0.6%) | 587 (3.2%) | 0.25 (0.12,0.45) | 9.69E-08 |
| Laurin-sandrow syndrome | Disease or Syndrome | 15 (1.0%) | 706 (3.9%) | 0.32 (0.18,0.51) | 1.23E-07 |
| Hispanics | Population Group | 30 (2.0%) | 71 (0.4%) | 3.66 (2.33,5.61) | 1.23E-07 |
| Fluid output.urine | Finding | 3 (0.2%) | 374 (2.1%) | 0.14 (0.04,0.34) | 1.51E-07 |
| Anemia; Unspecified | Disease or Syndrome | 26 (1.8%) | 964 (5.3%) | 0.40 (0.27,0.59) | 2.00E-07 |
| Patient Identity Verified (Finding) | Finding | 3 (0.2%) | 311 (1.7%) | 0.14 (0.04,0.35) | 2.21E-07 |
| Planning | Mental Process | 195 (13.1%) | 3636 (20.0%) | 0.68 (0.58,0.79) | 3.48E-07 |
| Pmh - Past Medical History | Finding | 80 (5.4%) | 1887 (10.4%) | 0.57 (0.45,0.72) | 4.55E-07 |
| Inspiration Function | Organism Function | 54 (3.6%) | 1420 (7.8%) | 0.52 (0.39,0.68) | 5.06E-07 |
| Sees hand movements (Finding) | Finding | 7 (0.5%) | 5 (0.0%) | 22.18 (7.26,71.96) | 5.60E-07 |
| Probable | Finding | 37 (2.5%) | 1114 (6.1%) | 0.47 (0.33,0.64) | 5.73E-07 |
| Dilatation | Finding | 16 (1.1%) | 691 (3.8%) | 0.35 (0.20,0.55) | 6.16E-07 |
| Convulsions | Sign or Symptom | 10 (0.7%) | 537 (2.9%) | 0.28 (0.14,0.50) | 7.28E-07 |
| Suspiciousness(negated) | Finding | 12 (0.8%) | 595 (3.3%) | 0.31 (0.17,0.52) | 7.50E-07 |
| Not Difficult at all | Finding | 9 (0.6%) | 501 (2.8%) | 0.27 (0.13,0.49) | 7.71E-07 |
| G1 Grade (Finding) | Finding | 2 (0.1%) | 309 (1.7%) | 0.12 (0.03,0.34) | 8.01E-07 |
| Disease, Metabolic | Disease or Syndrome | 17 (1.1%) | 746 (4.1%) | 0.36 (0.21,0.56) | 8.35E-07 |
| Pneumothorax(negated) | Disease or Syndrome | 161 (10.9%) | 3280 (18.0%) | 0.66 (0.55,0.78) | 8.87E-07 |
| Normal vital signs (Finding)(negated) | Finding | 34 (2.3%) | 146 (0.8%) | 2.88 (1.94,4.17) | 9.60E-07 |
| Pleural effusion Disorder(negated) | Disease or Syndrome | 127 (8.6%) | 2674 (14.7%) | 0.64 (0.53,0.77) | 9.94E-07 |
| Pneumonias, Viral | Disease or Syndrome | 33 (2.2%) | 180 (1.0%) | 2.88 (1.94,4.15) | 1.00E-06 |
| Problem | Finding | 191 (12.9%) | 3525 (19.4%) | 0.69 (0.58,0.80) | 1.15E-06 |
| Characteristic of Perceptual Performance (Observable Entity) | Mental Process | 5 (0.3%) | 382 (2.1%) | 0.21 (0.08,0.43) | 1.32E-06 |
| Seen on arrival (Finding) | Finding | 20 (1.3%) | 729 (4.0%) | 0.39 (0.24,0.60) | 2.17E-06 |
| Caucasians | Population Group | 7 (0.5%) | 421 (2.3%) | 0.25 (0.11,0.48) | 2.25E-06 |
| Patient position (Attribute)(negated) | Finding | 3 (0.2%) | 269 (1.5%) | 0.16 (0.04,0.39) | 2.25E-06 |
| Altered Mental Status (Finding) | Mental or Behavioral Dysfunction | 7 (0.5%) | 469 (2.6%) | 0.25 (0.11,0.48) | 2.46E-06 |
